# Supervised exercise training in patients with cancer during anthracycline-based chemotherapy to mitigate cardiotoxicity – a randomized controlled trial

**DOI:** 10.1101/2023.02.06.23285559

**Authors:** Caroline Schneider, Christoph Ryffel, Manuela Rabaglio, Thomas M. Suter, Kristin L. Campbell, Prisca Eser, Matthias Wilhelm

**Author notes:** Address for correspondence including e-mail address: Matthias Wilhelm, Medical Division of Rehabilitation & Sports Medicine, Inselspital, Bern University Hospital, University of Bern CH-3010 Bern, Switzerland.

## Abstract

**Background:** Exercise training (ET) has been shown to mitigate cardiotoxicity of anthracycline-based chemotherapies (AC) in animal models. Data from randomized controlled trials in patients with cancer are sparse.

**Methods:** Patients with breast cancer or lymphoma receiving AC were recruited from four cancer centres and randomly assigned to three months supervised ET during (EXduringAC) or after (EXpostAC) AC. All patients were counselled on physical activity (PA) and PA was objectively measured with an activity tracker with PA feedback option. Primary endpoint was change in left ventricular global longitudinal strain (GLS) between baseline and AC completion (AC-end) assessed by transthoracic 2D echocardiography. Secondary endpoints were change in high sensitivity Troponin T (hsTnT) and NT-pro-brain natriuretic peptide (NT-proBNP) between baseline and AC-end. Exploratory analyses were performed at 6 months (follow-up). Primary and exploratory analysis were performed by linear mixed models and robust linear models, respectively.

**Results:** One-hundred-and-forty-three patients were eligible for the study. Fifty-seven patients (median age 47 [1^st^ and 3^rd^ quartiles 38, 57] years; 95% women) consented to participate, of whom 28 and 29 patients were randomized to the EXduringAC and EXpostAC group, respectively. GLS deteriorated in both study groups from baseline to AC-end with 7.4% and 6.2% relative increase in EXduringAC and EXpostAC, respectively, and hsTnT and NT-proBNP significantly increased in both groups, without difference between groups for any parameter. Duration of moderate-to-vigorous PA (MVPA) was also similar between groups with a median of 33 [26, 47] min/day and 32 [21, 59] min/day in the EXduringAC and EXpostAC group, respectively. In the robust linear model including the pooled patient population, MVPA was significantly associated with a more negative GLS and lesser increase in hsTnT.

**Conclusion:** In physically active patients with cancer receiving AC, supervised ET during chemotherapy was not superior to PA advice and tracker monitoring only to mitigate cardiotoxicity. Objectively measured PA was inversely related to markers of LV function and myocardial injury.

**Clinical Perspective:** What is new?
- This is the first study to demonstrate a significant inverse association between objectively measured MVPA and markers of LV dysfunction and myocardial injury.
- Objectively measured PA revealed that in physically active patients with cancer, the addition of supervised ET did not result in more PA compared to PA advice and feedback by activity trackers alone.

*What are the clinical implications?:*

- In a physically active cohort, PA counselling combined with PA tracking represents a viable alternative compared to supervised ET to mitigate cardiotoxicity and should be offered to patients undergoing AC.
- Our findings suggest that there is a dose-response relationship between PA and cardioprotective effects during AC.

## Introduction

Due to improvements in anticancer therapies and early cancer detection, the number of cancer survivors has increased substantially over the last decades.^1^ There is a need for adequate long-term care of cancer survivors including the management of side-effects from cancer treatment. Anthracycline-based chemotherapies (AC) form an integral part of antineoplastic regimens in the treatment of breast cancer and lymphoma and have a dose-dependent adverse effect on the heart and/or vascular endothelial function.^2^ Although various mechanisms have been proposed to explain the AC-induced cardiovascular (CV) toxicity,^3^ the majority of studies suggest that an increase in oxidative stress, evidenced through the generation of reactive oxygen species and binding on topoisomerase-IIβ, may cause cardiac myocyte apoptosis, necrosis and damage to myocyte mitochondria.^4,5^ The presence of shared risk factors (i.e., inactive lifestyle, obesity, smoking)^6^ in conjunction with AC-induced CV toxicity further augment the risk of cardiovascular disease (CVD) in this population.^7^ Indeed, a recent study reports that breast cancer survivors have a 1.8-fold increased risk of death from CVD 7 years post-diagnosis compared to age-matched women from the general population with CVD accounting for 35% of non-breast-cancer related mortality.^8^

While left ventricular (LV) remodelling and contractile dysfunction may not be apparent until months to years after chemo- and/or radiotherapy,^9^ other markers enabling the detection of subclinical cardiotoxicity have been proposed. These markers include a decrease in LV global longitudinal strain (GLS) and elevation of biochemical markers of acute myocardial injury and haemodynamic stress, such as high-sensitivity Troponin-T (hsTnT) and N-terminal pro-brain natriuretic peptide (NT-proBNP), respectively.^10^

Exercise-based cardio-oncology rehabilitation programmes (CORE) have been recommended by the 2022 ESC Guidelines on cardio-oncology.^11^ While structured exercise training (ET) has been shown to prevent and/or recover loss in cardiorespiratory fitness (CRF)^12^ and to alleviate side-effects of cancer therapies (i.e., reduce fatigue and improve quality of life),^13,14^ it remains unknown whether ET during AC-therapy can prevent a decline in cardiac function.^15^

Evidence for a cardioprotective effect of ET on cardiac markers during AC therapy stems mainly from animal studies^16,17^ which have been summarized in two meta-analyses, suggesting that ET improves fractional shortening, a marker of systolic function, in trained rats compared to their sedentary counterparts.^18,19^ Both meta-analyses found greater cardioprotective effects when the ET programme was started prior to AC-therapy and conducted concomitantly to AC-therapy in comparison to after Doxorubicin exposure.^18,19^

In humans, data from cross-sectional and cohort studies assessing the association between self-reported physical activity (PA) and cardiac function support the hypothesis of a cardioprotective effect of exercise.^20,21^ However, randomized controlled trials in humans are sparse, and results from animal studies could not be fully translated.^22,23^

We therefore aimed to compare changes in markers of cardiac function and myocardial injury between patients performing supervised ET during AC and patients receiving only advice on PA, with all patients receiving an activity tracker. We hypothesized that ET during AC would mitigate the decline in GLS and increase in hsTnT and NTpro-BNP, compared to PA advice only.

## Methods

### Study design

The CAPRICE-Study (Cancer Adverse Effects Prevention with Care & Exercise) was a prospective, randomized controlled multicentre study, investigating the effect of exercise therapy timing with regard to cardiotoxicity and patient preference in early breast cancer and lymphoma patients receiving AC therapies (clinicaltrials.gov number NCT03850171). The study was conducted in four large Swiss cancer referral centres including one university hospital after it was approved by the Ethic Committee of the Canton of Bern, Switzerland.

Women and men aged 18 years and older with histologically confirmed breast cancer and lymphoma who were scheduled for first time AC were eligible for this study. Participants were excluded if they had: 1) a known or newly diagnosed structural heart disease; 2) a contraindication to maximal cardiopulmonary exercise testing (CPET); 3) cancer-specific contraindications to exercise; 4) previous radiotherapy of the mediastinum and/or the left breast; or 5) significant cognitive impairment or inability to follow the study procedures (i.e., due to language problems).

Group allocation was done using the minimization method,^24^ implemented in the randomization software MinimPy^25^ to balance between group characteristics as follows: type of cancer (breast cancer or lymphoma), age (< or ≥50 years, baseline left ventricular global longitudinal strain (≤-19% or >-19%), previous cancer therapy (previously treated with cardiotoxic therapy or not previously treated with cardiotoxic therapy), human epidermal growth factor receptor 2 (HER2) status (positive or negative) and chemotherapy cycle length (2 weeks or 3 weeks). The allocation was performed after recruitment by an independent study group at the University Hospital. The intervention group participated in a centre-based cardio-oncologic rehabilitation (CORE) programme during AC (EXduringAC) while the control group (EXpostAC) performed the CORE programme after AC. Breast cancer patients received 4 doses of AC either in a 14-day interval (dose-dense) or in a 21 day interval whereas lymphoma patients underwent 2-6 cycles of either R-CHOP or O-CHOP, escalated BEACOPP or ABVD-therapy with doxorubicin-administration every 3 weeks. A complete list depicting all administered AC is shown in the supplemental material (Table S1).

Patients attended the laboratory for three visits: before or within 2 weeks of AC-initiation (baseline, week −1 to 2), in the first two weeks upon AC completion (AC-end, week 8-14) and at 12 weeks after AC completion (follow-up, week 20-26). During all visits, patients provided a blood sample, completed an echocardiogram, a CPET for assessment of cardiorespiratory fitness (peak oxygen consumption) and a bio-impedance scan to obtain their body composition. Health-related quality of life (QoL) and fatigue were assessed using the validated functional assessment of chronical illness (FACIT) questionnaire, whereas physical activity pattern were evaluated with the global physical activity questionnaire (GPAQ). All patients received PA counselling at the baseline visit and were encouraged to perform leisure time PA according to current recommendations.^26^ They received a hip- or bra-worn Fitbit Zip PA tracker for the whole 6-month study duration and were instructed to wear it every day.

In this paper we report the primary outcome change in GLS from baseline to AC-end, and selected secondary outcome parameters at AC-end and follow-up (change in hsTnT, NT-proBNP). Detailed data on secondary outcomes (CRF, quality of life and fatigue) will be reported in a related paper.

### Supervised exercise training

The supervised ET intervention was embedded in a 12-week ambulatory CORE programme, including 24 supervised centre-based and 12 non-supervised home-based exercise sessions, counselling on PA, psychological aspects, nutrition, cardiovascular risk factors and pain management.^27^ The ET was offered twice weekly to groups of approximately 8 to 10 patients supervised by experienced exercise therapists. Sessions started with approximately 30-40 min of cycling on an ergometer at moderate intensity, increasing on a weekly basis, if possible. After the cycling training, patients continued with approximately 40-45 min of strength, coordination and/or balance training. The third weekly exercise session was performed at home and was supported by an activity tracking device with visual feedback on PA (see below).

### Adaptations of supervised ET during the Covid-19-pandemic

As a consequence of the Covid-19 related restrictions, CORE was adapted to a hybrid-tele-rehabilitation model, comprising of one supervised individual ET session and two non-supervised ET sessions at home.^28^ The home-based exercise sessions were supervised via the Fitbit Zip activity tracking device (Fitbit, San Francisco, USA) and feedback was provided by the therapist via telephone or at the training centre, if possible. During the Covid-19 pandemic, some patients completed a CORE without centre-based sessions. All patients were encouraged to perform at least 150 min of moderate PA per week, and two weekly strength sessions for major muscle groups.^26^

### Transthoracic echocardiography

Standard transthoracic 2D echocardiography was performed before randomisation, at AC-end and at follow-up. All echocardiographic images were obtained on a Vivid 95 cardiac ultrasound system with a 7.5-MHz transducer (GE Medical system, New Jersey, U.S.A.) by experienced sonographers of an echo core lab using standard tomographic views. All data was stored on an external hard-drive and analysed off-line on a commercially available workstation using TomTec software (TomTec Imaging Software Systems, 2020).

Traditional echocardiographic parameters of LV geometry, and systolic and diastolic function were assessed based on contemporary recommendations.^29,30^ LV end-diastolic (LVEDV), and LV end-systolic volumes (LVESV) were calculated using the biplane method. All volumes were indexed for body surface area (BSA). LV systolic function was expressed as ejection fraction (EF), derived from the LVEDV and LVESV. LV diastolic function was assessed by pulse-wave and tissue Doppler in the apical four-chamber view.

Peak systolic LV GLS was assessed using standard 2D apical four-chamber, two-chamber and three-chamber views using speckle-tracking analysis.^31^ Manual tracing of the endocardial borders on an end-systolic frame (aortic valve closure) was performed and the myocardial region of interest was adjusted to include all the endocardium and epicardium, excluding the pericardium. Automatic tracing was then applied on subsequent frames. Adequate tracing for each segment was verified and manually corrected, if necessary. GLS was determined by averaging all values of the 18 segments of the three views. LV GLS was assessed by an experienced echocardiographer blinded for group allocation and time point.

### Biomarkers of myocardial injury

Blood samples were obtained from the antecubital vein with the patient in a seated position at each visit preceding the CPET. We measured the plasma concentrations of hsTnT (in ng/L, Roche Elecsys, 2021, three centres), and NT-proBNP (in pg/ml, Roche Elecsys, all centres). One participating centre (5 patients) determined hs-Troponin-I instead of hsTnT. Due to non-comparability of these two biomarkers, only patients with available hsTnT were included in this analysis.

### Cardiopulmonary exercise testing

CPETs were performed on a cycle ergometer with an individualized ramp protocol aiming to achieve exhaustion within 8 to 12 min. The protocol consisted of a 3-min warm-up at a workload of 5-20 watt followed by an increase of 10, 15, or 20 watt per minute until voluntary exhaustion and a 2-min active cool-down period. Throughout the CPET, patients were monitored by a cardiologist with continuous assessment of a 12-channel electrocardiogram. From May 2019 until the end of February 2020 gas exchange was measured using the breath-by-breath spirometry system Jaeger Oxycon Pro (Masterscreen CPX, PanGas Healthcare GmbH, Dagmersellen, Switzerland) which was thereafter replaced by the Quark spirometric system (Cosmed, Fehraltdorf, Switzerland). Peak VO_2_ was determined as the highest value of a 30 s moving average window.

### Physical Activity Tracking

PA of all participants was tracked with a Fitbit Zip device, which was handed out to patients during baseline visit. Patients were asked to attach the device to their belt or bra and wear it all day every day until the follow-up visit, which amounted to a duration of 6 months for most patients. The device enabled constant recording of PA parameters such as daily steps and daily minutes of low, medium and high activity. Feedback regarding the daily steps could be seen by the patient on the screen of the device and greater detail on daily activity levels could be assessed by the patient with a login on the website. At the AC-end and follow-up visits data was downloaded from the Fitibit Zip by the study team. Since Fitbit does not provide information on the acceleration threshold used to distinguish between medium or high PA, data from these parameters were summed to obtain one parameter, which we termed moderate to vigorous activity (MVPA). Additionally, all participants kept a training diary in which they documented modality, duration and Borg Scale of all training sessions they completed, including individual exercise performed at home and exercise as part of the CORE programme. Training sessions not reflected by the Fitbit Zip, such as cycling or swimming, or when the Fitbit Zip device was not worn, were added to the PA data by a formula (see supplemental material).

Of the PA data from Fitbit Zip and training diary, the following parameters were calculated for each patient: daily steps including converted non-step based PA, and daily MVPA including converted non-step based PA.

### Serious adverse events

According to the Ordinance on Clinical Trials in Human Research, serious adverse events were defined as any untoward medical occurrence that I)results in death or is life-threatening, ii) requires in-patient hospitalisation or prolongation of existing hospitalisation or III) results in persistent or significant disability or incapacity. Serious adverse events were monitored by an independent data safety monitoring committee of the study and reported to the local ethics committee within 15 days.

### Sample size calculation

Sample size calculation was based on previous studies that measured GLS before and after AC in patients with breast cancer. Based on these studies, we assumed that the EXpostAC group will increase GLS by 2% from −20% to −18% and the EXduringAC group will have an increase of only 0.5% from −20% to −19.5% (with a standard deviation of 2.5%). Based on a one-sided t-test, an alpha of 0.05 and power of 0.8, 102 patients (51 patients per group) were required, assuming a 15% dropout rate.

### Data analysis

We used the R Studio software (Version 2022.02.3+492, R version 4.2.1.) for all statistical analyses.

Patient characteristics were compared between groups using Wilcoxon rank sum tests, Chi-square or Fisher’s exact test, as appropriate.

The effect of the exercise intervention on changes in markers of LV function and myocardial injury were compared between the two groups by linear mixed model with group (EXduringAC and EXpostAC) and time point (visits) as fixed factors including their interaction. Patients were included as random intercepts. Mixed models were used because they can handle missing at random data.^32^ Primary outcome was the group × time interaction for GLS from baseline to AC-end of patients with available GLS at both time points. An additional model was performed for GLS with heart rate as a covariate to adjust for higher heart rate that can arise as a consequence of AC-induced anaemia and consequent changes in preload. Due to assay-specific non-detection values of <5 ng/L and <50 pg/ml for hsTnT and NT-proBNP, respectively, a conservative approach was used in the linear mixed model by setting these values at 4 ng/L for hsTnT and at 49 pg/ml for NT-proBNP. Log transformation was used when necessary.

In exploratory analyses, robust linear models were performed for change in GLS, LVEDVi, hsTnT and NT-proBNP from baseline to AC-end for the pooled patient population, including treatment group and objectively measured PA (expressed as daily MVPA or steps) as predictor variables.

All analyses were performed on an intention-to-treat (ITT) basis. In addition, per protocol (PP) analyses were performed (Supplement Table S2). PP was defined according to which phase (AC or post-AC phase) patients performed at least 60% of the centre-based training sessions in. Patients who did not complete 60% of the planned training sessions in neither phase were excluded from the PP analysis. Missing data was not imputed.

## Results

### Study Flow and baseline characteristics

The patient flow is shown in Figure 1 (and for PP-analysis in Supplement Figure S1). Between May 2019 and June 2022, 143 patients were assessed for eligibility at four centres, and 57 patients consented to participate in the study and were randomized. Twenty-eight patients were allocated to EXduringAC and 29 to EXpostAC. Baseline demographic and clinical characteristics are shown in Table 1.

**Table 1:**
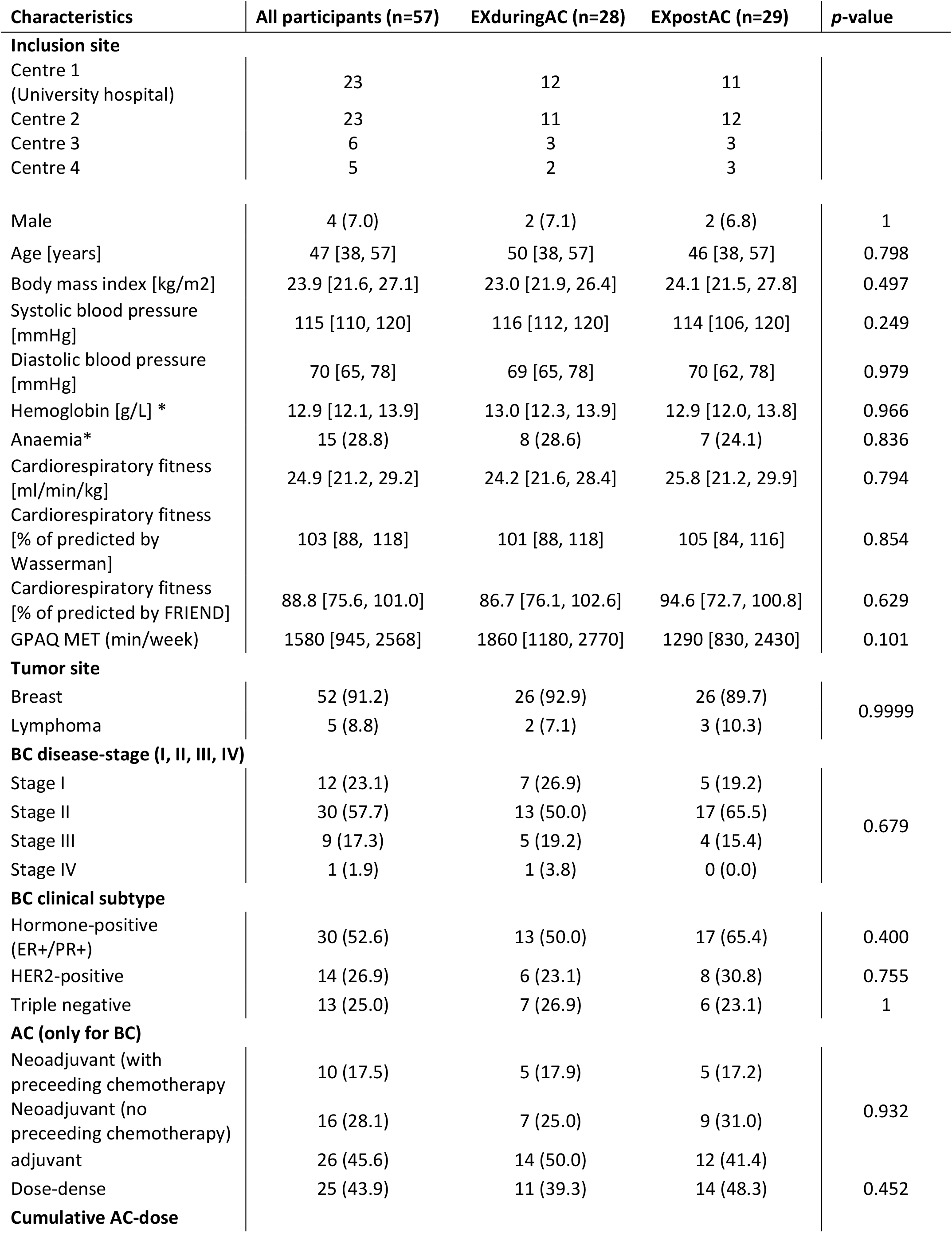
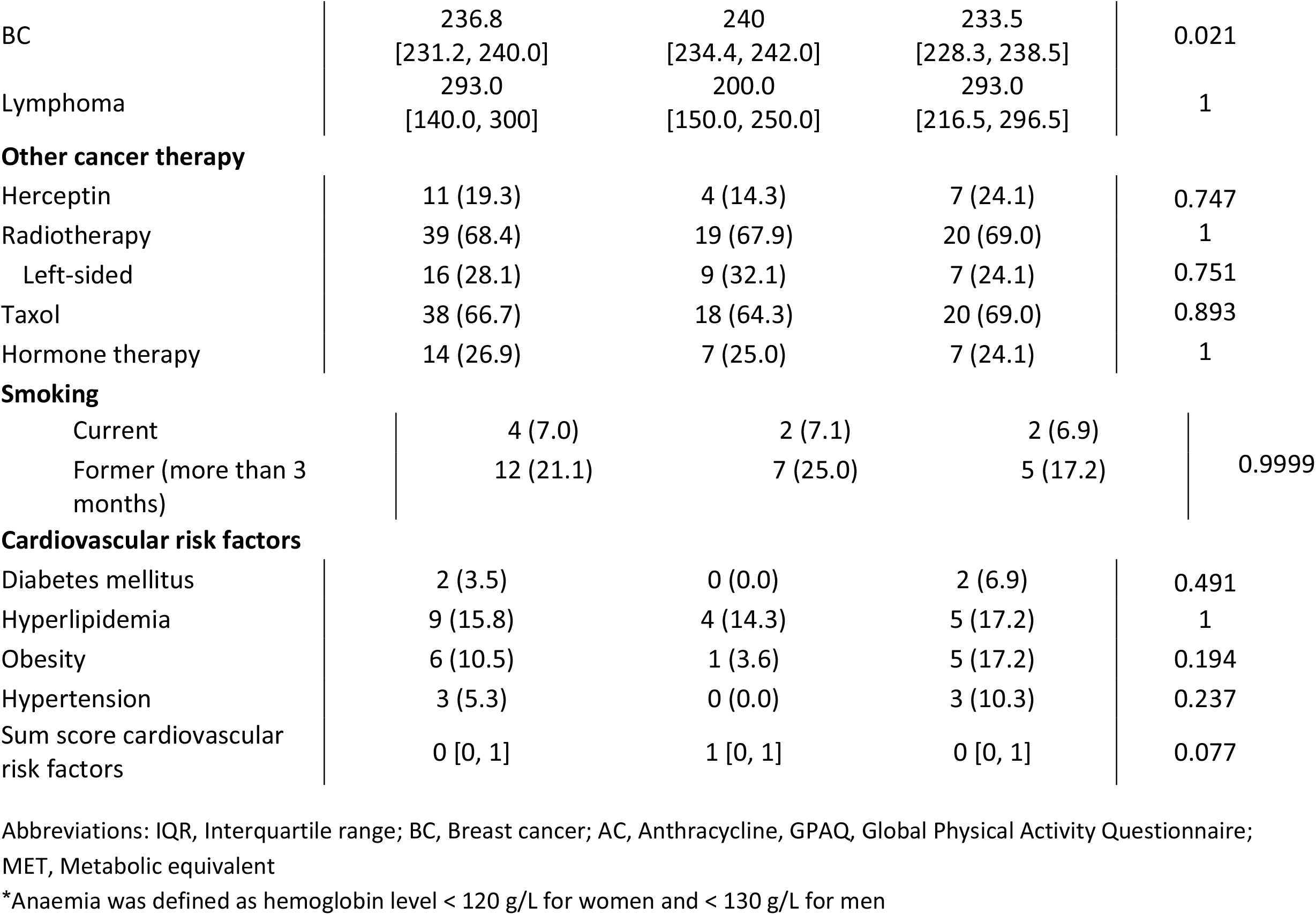
Baseline characteristics of the study population, shown are n (%), median [1^st^ and 3^rd^ quartiles]. Indicated are p-values from Wilcoxon two-sample tests or Fisher’s exact test, as appropriate.

**Figure 1:**
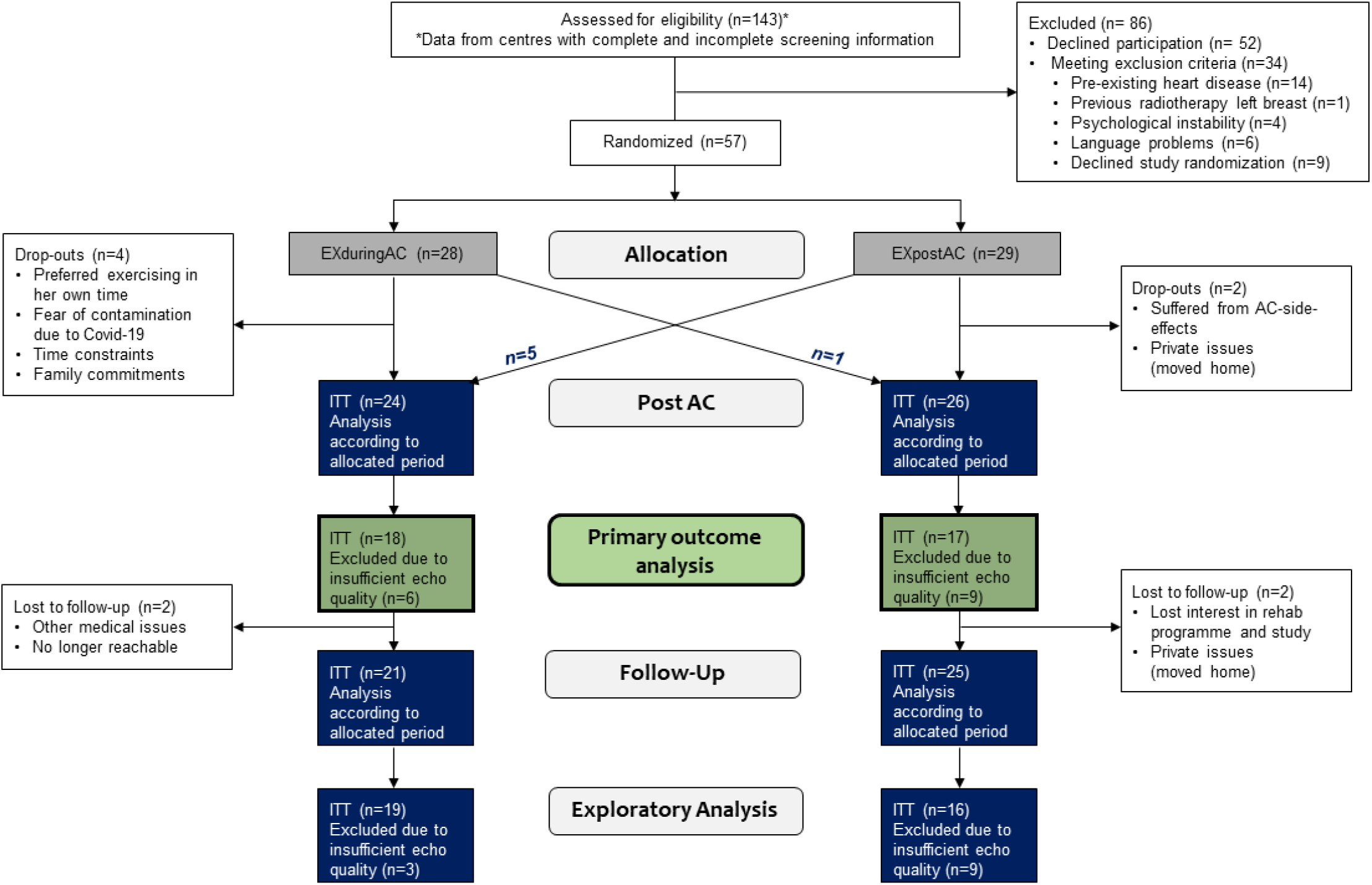
Study flow

Six patients (10.5%) discontinued the study before the AC-end visit (4 EXduringAC, 2 EXpostAC). Patients who dropped out of the study were 11 years older compared to the remaining study cohort (p=0.007) but did not differ with regard to other baseline characteristics (Supplement Table S2). Reasons for discontinuation of the study were time constraints due to private commitments and other medical appointments, side-effects from AC-therapy, or no longer being interested in the study. Six patients chose the timing of their CORE participation discordant with their randomised group allocation. However, results of all models were consistent between ITT and PP analyses, so the effect of non-compliant patients or patients changing group was negligible. Due to insufficient image quality at baseline and/or at AC-end and a missing visit related to Covid-19, primary outcome data of GLS could not be analysed in 16 patients.

Ninety-one percent of the participants had breast cancer and 9% had lymphoma. The majority of patients with breast cancer were diagnosed with HER2+ (53%) and 50% of breast cancer patients were undergoing neoadjuvant AC. Of these patients, 9 participants (4 EXduringAC, 5 EXpostAC) received additional (cardiotoxic) chemotherapy prior to AC administration, whereby three patients (2 EXduringAC, 1 EXpostAC) received 4 cycles of Carboplatin and 6 patients (4 EXpostAC, 2 EXduringAC-group) received 4 cycles of Trastuzumab prior to AC-therapy.

AC for all but one breast cancer participants consisted of 4 cycles with 60 mg/m^2^ doxorubicin combined with 600 mg/m^2^ cyclophosphamide. One patient not receiving doxorubicin was administered an equivalent dose of epirubicin. Median [1^st^ and 3^rd^ quartiles] cumulative dose in the EXduringAC group and EXpostAC group was 240 mg/m^2^ [234.4, 242.0 mg/m^2^] and 233.5 mg/m^2^ [228.3, 238.5 mg/m^2^], respectively, which was a small but significant difference between groups.

Two lymphoma patients were diagnosed with Hodgkin’s lymphoma (stages II and III), two patients suffered from diffuse large B-cell-lymphoma (stages II and III) and one patient was diagnosed with follicular lymphoma (stage III-IV). Three lymphoma patients received six cycles of either R-CHOP or O-CHOP-therapy, 1 patient received 4 cycles of escalated BEACOPP and 1 patient was treated with 2 cycles of ABVD-therapy. All participants were administered the prescribed AC-dose with no dose reductions occurring.

In one patient randomized to the EXduringAC group, LVEF dropped from 55% at baseline to 39% at follow-up (−29%), which we documented as a serious adverse event. After evaluation with the data safety monitoring committee of the study, it was concluded that a causal relation of the event with the study intervention was unlikely. The patient received treatment with an angiotensin-converting-enzyme inhibitor and beta-blocker and was thereafter regularly monitored in the cardio-oncology-unit of the University Hospital.

### Intervention Adherence

Median [1^st^, 3^rd^ quartile] adherence with the centre-based exercise sessions over the 12-week CORE was 75% [49, 89%], with adherence in the EXpostAC group being 79% [71, 92%] vs. 71% [29, 83%] in the EXduringAC group.

### Changes in echocardiographic parameters and biomarkers

Baseline data and changes in echocardiographic parameters are shown in Table 2. Individual changes in GLS, LVEDVi, hsTnT and NT-proBNP are shown in Supplement Figure S2.

**Table 2:** Changes of echocardiographic parameters and biomarkers of myocardial injury according to allocated study group (Intention-to-treat analysis). Shown are median (1^st^ and 3^rd^ quartiles). Significant p-values for group and time interaction or main effects are indicated and derived from mixed linear models with patients as random factors (intercepts).

|  | EXduringAC (n=28) |  |  | EXpostAC (n=29) |  |  |
| --- | --- | --- | --- | --- | --- | --- |
|  | Baseline | Δ baseline to AC-end* | Δ AC-end to follow-up | Baseline | Δ baseline to AC-end* | Δ AC-end to follow-up |
| GLS [%] | -21.9<br>(-22.8, -19.1)<br>N=24 | -1.05<br>(-2.9, 3.1)<br>N=18 | 0.6<br>(-1.1, 2.7)<br>N=19 | -21.05<br>(-22.8, -19.1)<br>N=24 | 0.5<br>(-1.3, 2.6)<br>N= 17 | -1.1<br>(-3.2, 3.8)<br>N= 16 |
| HR GLS [bpm] | 71.5<br>(67.0, 79.8)<br>N=24 | 0.5<sup>b</sup><br>(-5.8, 12.3)<br>N=18 | -5.0<br>(-8.8, 1.5)<br>N=18 | 67.0<br>(62.5, 76.3)<br>N=24 | 8.5<sup>b</sup><br>(3.0, 14.3)<br>N=18 | -3.5<br>(-8.0, 2.5)<br>N=16 |
| 2D LVEF [%] | 64.5 (57.7, 68.3)<br>N=26 | -1.5 (-5.7, 3.4)<sup>b</sup><br>N=22 | 0.2 (-5.9, 4.4)<br>N=21 | 63.9 (61.4, 67.9)<br>N=29 | -3.5 (-6.0, 0.3)<sup>b</sup><br>N=25 | 2.0 (-3.9, 4.0)<br>N=23 |
| LVEDVi [ml/m<sup>2</sup>] | 52.3 (43.8, 60.6)<br>n=25 | -2.6 (-6.6, 4.4)<br>N= 21 | -5.8 (-10.2, 7.9)<br>N=21 | 47.3 (39.0, 53.1)<br>N=26 | 0.8 (-9.5, 7.7)<br>N=24 | -0.4 (-13.1, 6.2)<br>N=23 |
| E/e' | 6.6 (6.1, 8.1)<br>N=28 | -0.04 (-1.55, 0.91)<br>N=24 | 0.47 (-1.37, 1.00)<br>N=22 | 7.2 (5.7, 8.1)<br>N=29 | -0.36 (-0.78, 1.49)<br>N=26 | 0.00 (-0.90, 0.84)<br>N=23 |
| E/A ratio | 1.12 (1.0, 1.37)<br>N=28 | -0.09 (-0.22, 0.08)<br>N=25 | 0.00 (-0.20, 0.24)<br>N= 23 | 1.25 (0.93, 1.44)<br>N=29 | -0.09 (-0.14, 0.20)<br>N=26 | -0.06 (-0.37, 0.12)<br>N=24 |
| LAVI [ml/m<sup>2</sup>] | 22.6 (19.9, 28.4)<br>N=26 | -2.7 (-4.6, 3.11)<br>N=22 | 0.3 (-3.2, 5.9)<br>N=21 | 20.6 (18.9, 26.7)<br>N=29 | 0.4 (-1.3, 3.7)<br>N=23 | -0.9 (-4.3, 2.2)<br>N=21 |
| NT-proBNP [pg/ml] | 56.5 (49.0, 82.0)<br>N=28 | 35.0 (0.0, 107.0)<sup>b</sup><br>N=21 | -1.0 (-71.0, 41.0)<br>N=19 | 49.0 (49.0, 53.3)<br>N=28 | 41.0 (0.0, 93.0)<sup>b</sup><br>N=23 | -37.0 (-107.0, 0.0)<br>N=21 |
| hsTnT [ng/L] | 4.0 (4.0, 4.0)<br>N=25 | 8.0 (6.0, 16.0)<sup>b</sup><br>N=21 | -2.0 (-10.0, 1.0)<sup>b</sup><br>N=18 | 4.0 (4.0, 4.0)<br>N=25 | 10.0 (6.3, 14.8)<sup>b</sup><br>N=22 | -5.0 (-9.0, -1.0)<sup>b</sup><br>N=21 |
| hs-C-Reactive Protein [mg/l] | 1.03 (0.77, 1.58)<br>N=25 | 0.98 (-0.06, 2.49)<br>N=22 | -0.96 (-2.63, 0.29)<br>N=18 | 1.91 (0.97, 3.68)<br>N=26 | 0.81 (-0.02, 1.37)<br>N=22 | -0.56 (-2.83, 0.73)<br>N=22 |
| Systolic BP [mmHg] | 122.5<br>(116.0, 134.5)<br>N=26 | -5.5 (-13.0, 10.0)<br>N=20 | 1.0 (-10.0, 15.0)<br>N=17 | 120.0<br>(111.5, 135.5)<br>N=27 | 0.5 (-8.8, 14.8)<br>N=22 | -2.0 (-5.0, 5.5)<br>N=20 |
| Diastolic BP [mmHg] | 75.0 (65.8, 84.8)<br>N=26 | -2.0 (-10.3, 7.0)<br>N=20 | -3.0 (-13.0, 12.0)<br>N=17 | 74.0 (63.0, 80.0)<br>N=27 | -1.5 (-9.5, 11.3)<br>N=22 | 1.0 (-6.3, 9.3)<br>N=20 |

a, $p < 0.05$ between groups at baseline; b, significant main effect for time; c, significant interaction effect for study group and time; \*primary outcome visit. Reference group was ExpostAC.
Abbreviations: GLS, global longitudinal strain; HR, heart rate; LVEF, left ventricular ejection fraction; LVEDVi, left-ventricular end-diastolic volume index; hsTnT, high-sensitivity Troponin-T; NT-proBNP, N-terminal pro-brain natriuretic peptide; LAVI, left atrial end-systolic volume index; hs-C-Reactive Protein, high-sensitivity C-Reactive Protein; BP, Blood pressure.

At baseline, echocardiographic and biomarkers were not significantly different between groups.

Changes from baseline to AC-end in echocardiographic markers were similar with no differences between the two study groups and consistent results in the ITT and PP analyses (Table 3 and Supplement Table S3).

**Table 3:** Mixed linear models, including group x time interaction and the following predictor variables: studygroup. Reference category was EXpostAC. Analysis was done on an Intention-to-treat-basis.

|  | Estimate | Standard<br>error | t-value | p-value |
| --- | --- | --- | --- | --- |
| **GLS** |  |  |  |  |
| Intercept | -21.09 | 0.63 | -33.71 | 0.000 |
| EXduringAC | 0.46 | 0.88 | 0.52 | 0.605 |
| AC-end | 0.76 | 0.77 | 0.99 | 0.326 |
| Follow-up | 0.83 | 0.73 | 1.14 | 0.257 |
| EXduringAC x Visit 2 | -0.28 | 1.07 | -0.26 | 0.792 |
| EXduringAC x Visit 3 | 0.93 | 1.05 | 0.88 | 0.381 |
| **LVEDVi** |  |  |  |  |
| Intercept | 46.77 | 2.44 | 19.20 | 0.000 |
| EXduringAC | 4.95 | 3.47 | 1.43 | 0.159 |
| AC-end | 0.20 | 2.59 | 0.08 | 0.938 |
| Follow-up | -0.74 | 2.66 | -0.28 | 0.781 |
| EXduringAC x Visit 2 | -0.42 | 3.75 | -0.11 | 0.910 |
| EXduringAC x Visit 3 | 0.80 | 3.89 | 0.20 | 0.839 |
| **Log hsTnT** |  |  |  |  |
| Intercept | 1.42 | 0.09 | 16.49 | 0.000 |
| EXduringAC | -0.01 | 0.12 | -0.05 | 0.961 |
| AC-end | 1.21 | 0.11 | 10.68 | **0.000** |
| Follow-up | 0.76 | 0.12 | 6.63 | **0.000** |
| EXduringAC x Visit 2 | 0.01 | 0.16 | 0.06 | 0.954 |
| EXduringAC x Visit 3 | 0.19 | 0.17 | 1.12 | 0.268 |
| **Log NT-proBNP** |  |  |  |  |
| Intercept | 4.03 | 0.11 | 35.62 | 0.000 |
| EXduringAC | 0.13 | 0.16 | 0.78 | 0.438 |
| AC-end | 0.57 | 0.13 | 4.21 | **0.000** |
| Follow-up | 0.17 | 0.14 | 1.23 | 0.223 |
| EXduringAC x Visit 2 | -0.13 | 0.19 | -0.66 | 0.514 |
| EXduringAC x Visit 3 | 0.20 | 0.20 | 0.98 | 0.332 |

Abbreviations: GLS, global longitudinal strain, LVEDVi, left-ventricular end-diastolic volume index, hsTnT, high-sensitivity Troponin-T, NT-proBNP, N-terminal pro-brain natriuretic peptide

GLS at baseline was within the age-expected range. In patients with available GLS at baseline and AC-end, GLS increased and hence deteriorated from −21.6% [-22.3, −19.0%] to −20.0% [-22.7, −18.8%] in the EXduringAC group (7.4% relative increase) and from −21.1% [-22.6, −18.9%] to −19.8% [-23.8, −16.9%] in the EXpostAC group (6.2% relative increase, Figure 2).

**Figure 2:**
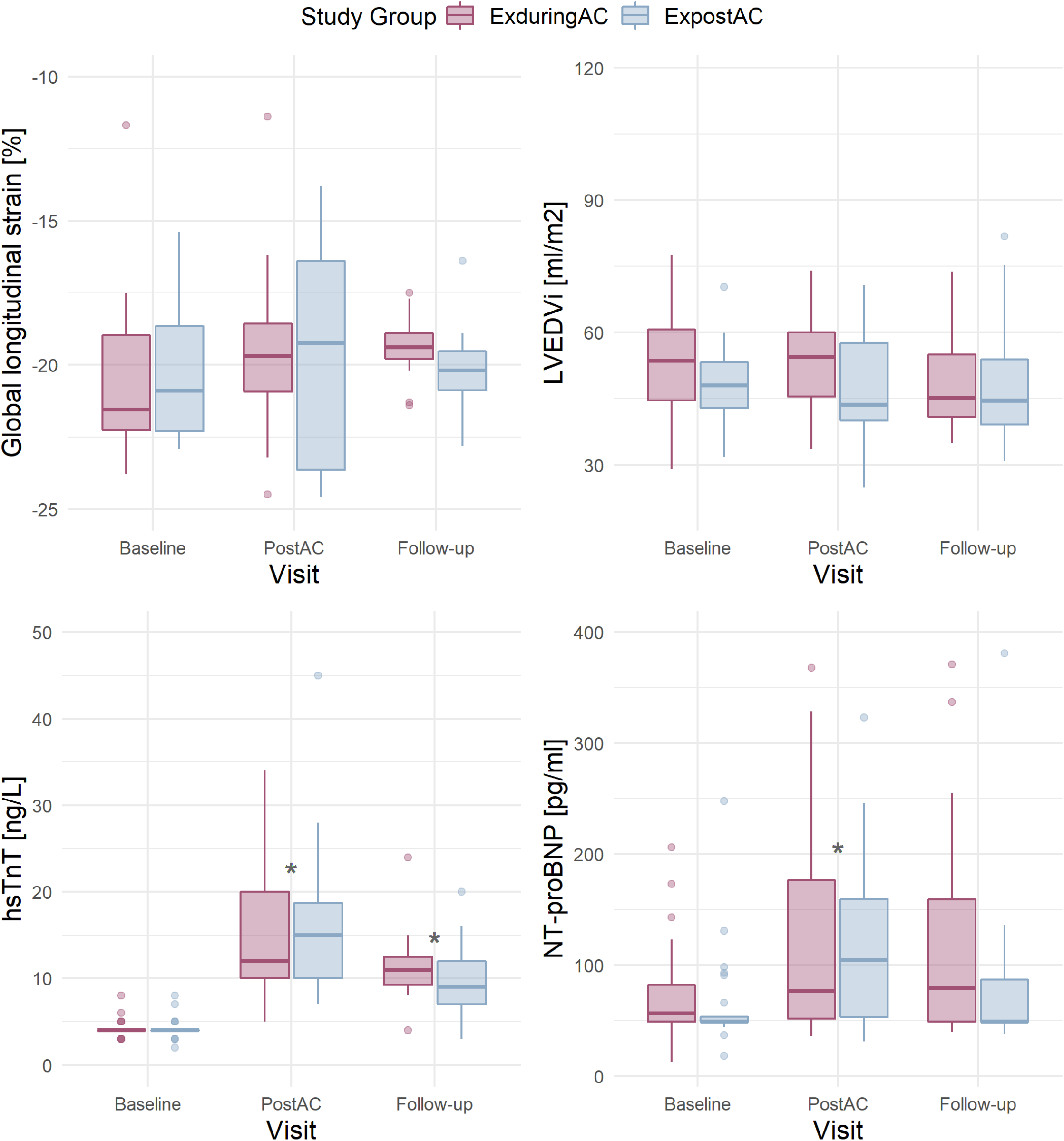
Boxplots per study group (according to Intention-to-treat) and visit in patients with available data at baseline and AC-end for GLS (n= 18 in the EXduringAC and n= 17 in the EXpostAC group), LVEDVi (n= 21 and n=24), hsTnT (n= 21 and n=22) and NT-proBNP (n=24 and n= 25). AC, anthracycline-based chemotherapies; GLS, global longitudinal strain; LVEDVi, left-ventricular end-diastolic volume index; hsTnT, high-sensitivity Troponin-T; NT-proBNP, N-terminal pro-brain natriuretic peptide

At no time points did the change in GLS exceed the threshold of 15% that has been suggested to predict a future drop in LVEF.^33^ In an additional linear mixed model adjusted for heart rate, a higher heart rate was associated with a less negative GLS, whereby an increase in heart rate of 10/min resulted in an 0.8% change in GLS (Supplement Table S4).

At AC-end, there was a significant increase in hsTnT by a median of 8.0 ng/L [6.0, 16.0 ng/L] and 10.0 ng/L [6.3, 14.8 ng/L] in the EXduringAC and EXpostAC group, respectively. NT-proBNP also increased significantly by a median of 35.0 pg/ml [0.0, 107.0 pg/ml] and 41.0 pg/ml [0.0, 93.0 pg/ml] in the EXduringAC and EXpostAC group, respectively, with no changes between the groups (p for interaction group x time = 0.954 and 0.514 for hsTnT and NT-proBNP, respectively). The increase in hsTnT persisted at follow-up, whereas NT-proBNP was no longer different when compared to baseline (Table 2, Figure 2).

### Physical activity data

At baseline the overall volume and intensity of PA did not differ between EXduringAC and EXafterAC, except for a lower step count during AC cycle 1 in ExduringAC compared to ExpostAC. Moderate-to-vigorous PA averaged to a median of 33 min/day [26, 47] min/day and 32 min/day [21, 59] min/day in the EXduringAC and EXpostAC group, respectively. This corresponded to an average of 7641 steps/day [6332, 9810] steps/day in the EXduringAC group and 6806 steps/day [5275, 9429] steps/day in the EXpostAC group. Overall, 76% of our cohort reached the recommended amount of MVPA of 150 min/week during AC.

### Association of physical activity with markers of LV dysfunction and myocardial injury at AC-end

In the linear models including the pooled patient population, a higher step count was associated with a more negative GLS, with an increase of 2000 steps/day corresponding to a 1% more negative GLS. A similar association was found for MVPA with GLS and hsTnT, with a 30 min increase in MVPA corresponding to a 1.8% more negative GLS and a 3 ng/L lesser increase in hsTnT. We did not find any associations between activity parameters and LVEDVi or NT-proBNP (Table 4 and Figure 3).

**Table 4:** Robust linear models for changes in GLS, LVEDVi, hsTnT and NT-proBNP from baseline to AC-end, including study group and activity parameters as predictors. For GLS, models were adjusted for heart rate. Reference category was EXpostAC. Group allocation was according to Intention-to-treat. The models explained the following percentages of total variance for the model including total steps [min/day] and MVPA [min/day], respectively: 11% and 10% for peak GLS, 5% and 4% for LVEDVi, 3% and 6% for hsTnT and 1% and 2% for Nt-pro-BNP. In the models using total steps and MVPA as predictor, the final models are based on 28 and 27 patients for GLS, 36 and 35 patients for LVEDVi, 33 and 32 patients for hsTnT, and 34 and 33 patients for NT-proBNP, respectively.

|  | Estimate | Standard error | t-value | p-value |  | Estimate | Standard error | t-value | p-value |
| --- | --- | --- | --- | --- | --- | --- | --- | --- | --- |
| **GLS** |  |  |  |  | **GLS** |  |  |  |  |
| Intercept | 3.96 | 1.96 | 2.02 | 0.053 | Intercept | 3.03 | 1.44 | 2.10 | 0.045 |
| EXduringAC | -0.18 | 1.23 | -0.15 | 0.885 | EXduringAC | -0.62 | 1.20 | -0.51 | 0.611 |
| HR GLS | 0.07 | 0.08 | 0.83 | 0.412 | HR GLS | 0.02 | 0.06 | 0.32 | 0.755 |
| Steps | -0.0005 | 0.00 | -2.36 | **0.026** | MVPA | -0.06 | 0.03 | -2.30 | **0.030** |
| **LVEDVi** |  |  |  |  | **LVEDVi** |  |  |  |  |
| Intercept | -0.28 | 5.32 | -0.05 | 0.958 | Intercept | -2.44 | 4.51 | -0.54 | 0.592 |
| EXduringAC | -0.63 | 3.92 | -0.16 | 0.873 | EXduringAC | 0.05 | 3.94 | 0.01 | 0.989 |
| Steps | 0.00 | 0.00 | 0.04 | 0.971 | MVPA | 0.06 | 0.08 | 0.73 | 0.471 |
| **hsTnT** |  |  |  |  | **hsTnT** |  |  |  |  |
| Intercept | 19.21 | 4.37 | 4.40 | 0.000 | Intercept | 15.65 | 3.57 | 4.38 | 0.000 |
| EXduringAC | -1.19 | 2.94 | -0.41 | 0.688 | EXduringAC | -0.44 | 2.55 | -0.18 | 0.863 |
| Total steps | 0.00 | 0.00 | -1.71 | 0.097 | MVPA | -0.10 | 0.04 | -2.36 | **0.025** |
| **NT-proBNP** |  |  |  |  | **NT-proBNP** |  |  |  |  |
| Intercept | 85.57 | 28.61 | 2.99 | 0.005 | Intercept | 69.34 | 22.15 | 3.13 | 0.004 |
| EXduringAC | 6.18 | 26.73 | 0.23 | 0.819 | EXduringAC | 8.53 | 29.00 | 0.29 | 0.770 |
| Total steps | -0.01 | 0.00 | -1.67 | 0.104 | MVPA | -0.55 | 0.38 | -1.43 | 0.163 |

Abbreviations: MVPA, Moderate to vigorous physical activity; HR, heart rate, GLS, global longitudinal strain, LVEDVi, left-ventricular end-diastolic volume index, hsTnT, high-sensitivity Troponin-T, NT-proBNP, N-terminal pro-brain natriuretic peptide

**Figure 3:**
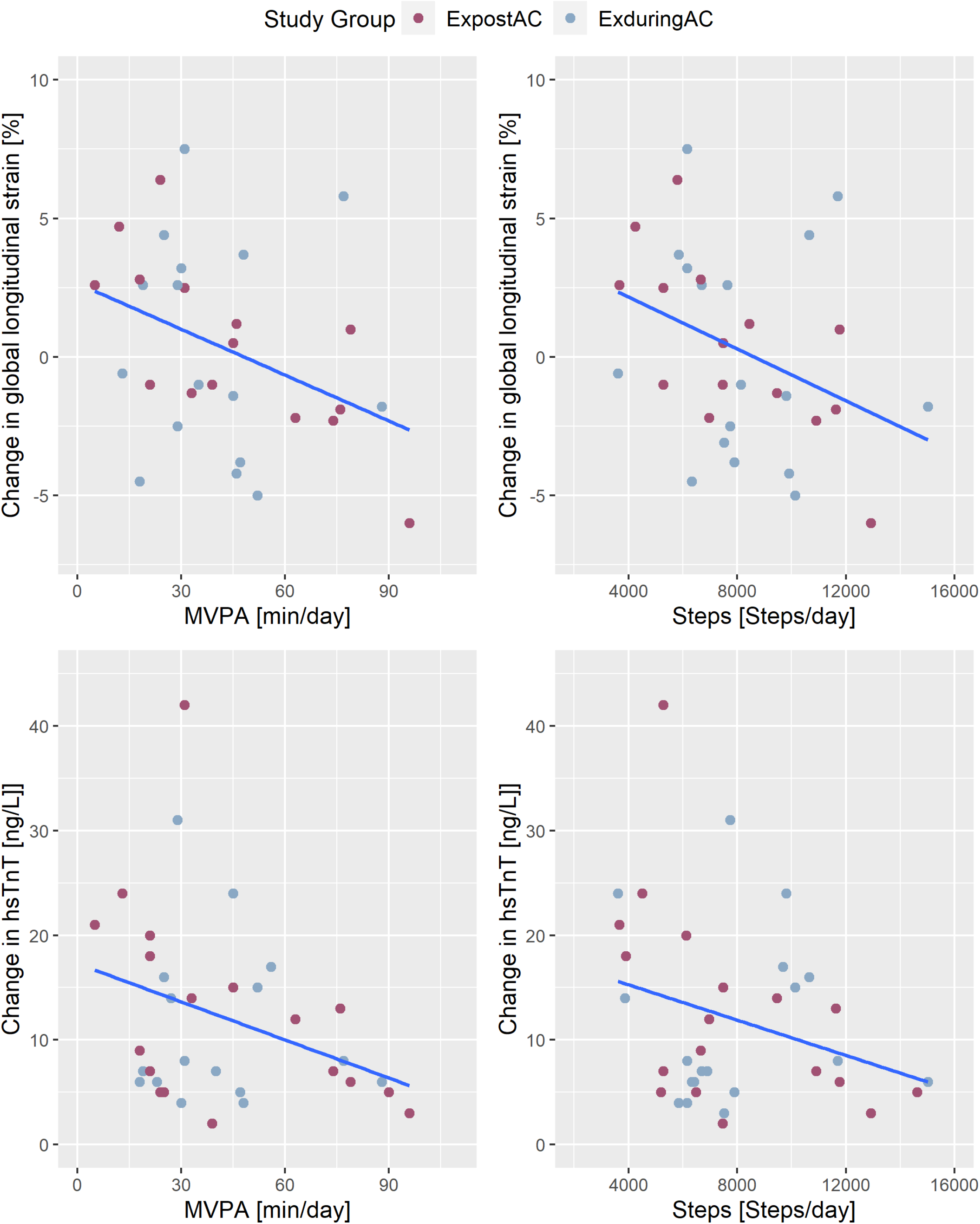
Association of activity parameters (MVPA and steps) with changes in GLS and hsTnT. MVPA, Moderate to vigorous physical activity; GLS, global longitudinal strain; hsTnT, high-sensitivity Troponin-T

## Discussion

In this randomized controlled trial in patients with cancer receiving AC, the addition of supervised ET during AC was not superior to PA advice and tracking alone to mitigate cardiotoxicity. Changes in GLS, hsTnT and NT-proBNP did not differ between the EXduringAC and EXafterAC groups. Importantly, objectively measured PA at baseline and AC-end achieved recommended levels of guidelines in both groups and did also not differ between the groups. In the pooled patient population, we found a significant inverse association between PA and changes in GLS and hsTnT. Given the consistency and evidence from experimental animal studies, the temporality of the association,^22^ the biological gradient, plausibility and coherence, this may suggest a causal association between PA and protection from AC cardiotoxicity in humans.^34^

Importantly, this is the first study using objectively measured PA with an activity tracker to analyse the association between overall PA and markers of myocardial function and myocardial injury. A recent study by Foulkes et al. in 104 breast cancer patients (mean age 51 years) undergoing AC compared cardiac markers of systolic function and biomarkers of myocardial injury between an exercise and usual care group.^35^ ET consisted of 3 weekly 30-60 min aerobic exercise sessions at moderate to high intensity, combined with resistance training over 12 weeks. In their study, GLS deteriorated by 1.3% in the exercise group compared to 1.0% in the usual care group with no differences between the groups. Another, non-randomized, study by Costell et al. assessed cardiac function using CMR in 27 early breast cancer patients at baseline and 3 weeks after AC completion.^36^ After the baseline visit, patients could choose whether they wanted to participate in an exercise intervention (3/week, 60 min/session) or be part of the usual care group. At completion of AC, a significant deterioration in GLS was found for the control group (1.6±1.4%, p=0.002) whereas GLS was preserved in the exercise group (−0.56±1.5%, p=0.377). However, no information was given on baseline characteristics and PA, and no causal associations could be implied due to the non-randomized study design. A recent prospective, observational study by Peck et al. assessed self-reported MVPA and cardiac function in 88 breast cancer patients over the course of AC- and HER2-therapy.^20^ In their study, every 30 min increase in MVPA during treatment was associated with a 0.04% improvement in GLS.

While absolute changes of the two studies that assessed GLS at baseline and AC-end are similar to our measurements,^35,37^ no information was given on overall PA of the included patients. Although Peck et al. analysed the relationship between MVPA and GLS and reported a significant association,^20^ the authors relied on self-reported measures which are likely to overestimate time spend at moderate to vigorous intensities.^38,39^

An increase in cardiac troponins (troponin-I, -T or hsTnT/I) upon AC has been described in numerous studies^37,40-42^ including a recent meta-analysis in 1048 patients which reported an odds ratio of 17.5 (95% CI 10.1-30.2) for an AC-induced increase in troponin levels.^43^ Similarly to results from our study, Foulkes and colleagues reported a significant increase in Troponin-I in both groups, which could not be prevented in the exercise group. However, their increase was significantly lower in the exercise group compared to the usual care group (8-fold vs. 16-fold baseline values),^35^ supporting our findings from the linear model in our pooled data showing an inverse association between exercise dose and troponin release. The same research group assessed BNP and Troponin-I in an earlier study with smaller sample size (28 patients) and found significant elevations in Troponin-I, but only a trend for a lesser increase in the exercise group (group x time p=0.10).^44^ In contrast to our study, they did not observe a significant increase in BNP. There is less evidence on the utility of NT-proBNP or BNP for both, prediction of cancer therapy-induced LV dysfunction^43^ and effects of exercise on the increase of this biomarker.^19^ Nevertheless, a seminal study by Kirkham and colleagues showed that a single treadmill session reduced the NT-proBNP elevation in the 24 to 48 hours after the first AC-cycle.^42^ When these single exercise bouts were completed before every AC cycle, however, a consistent reduction of the rise in NTproBNP or troponin by exercise could not be shown.^22,23^

The mechanisms leading to a rise in biomarkers following AC are not fully understood^45,46^ and were outside the scope of this study. In addition to acute cardiomyocyte injury, a rise in troponin may have been stimulated by cancer or cancer (-therapy) induced comorbidities such as anaemia or by certain hormones (i.e., catecholamines as a consequence of emotional stress).^45,46^ The association of a 30 min increase in total MVPA with a 3 ng/L lesser increase in troponin is encouraging in that it suggests that exercise may have a cardioprotective role.

A recent meta-analysis by Naaktgeboren et al. summarized the evidence for mechanisms responsible for ET-induced cardioprotection, which is mainly based on preclinical studies.^19^ Results from this analysis suggest that ET performed concomitantly to AC leads to an increased doxorubicin clearance and consequently less doxorubicin accumulation in cardiomyocytes, thereby inhibiting downstream pathways that may induce cardiotoxicity. Another hypothesis is the reduction of AC-induced oxidative stress (through upregulation of antioxidants and heat-shock-proteins) and the release of neuregulin^47^ with ET.

Against our hypothesis, ET participation during AC did not result in higher PA in the EXduringAC group. The fact that MVPA and steps were similar between the study groups allowed the pooled analysis including all patients in a linear model. Our findings can be explained by biologically plausible mechanisms and experimentally demonstrated results from animal studies.^19^ The dose-response relationship found in our and a self-reported previous study^20^ are suggestive of a causal relationship. Nevertheless, larger studies are needed to confirm our results and provide consistent evidence.

The significant increase in cardiac troponins could not be prevented by PA in any clinical study to date. Consequently, even though outcomes from preclinical studies and our findings suggest that PA during AC may have a cardioprotective effect, it remains unclear whether LV dysfunction can be prevented in the long-term. First evidence suggesting that protective effects may be lasting comes from the study by Foulkes and colleagues.^35^ In addition to resting cardiac parameters, this study compared measures of peak exercise cardiac function and reserve (cardiac output, stroke volume and LVEF) between the exercise and usual care group at the end of AC and after 12 months. They observed a better myocardial function and greater cardiac reserve in the exercise group compared to the usual care group at AC-end and these differences persisted at the follow-up assessment (in adherent patients).^35^ On the other hand, no differences were reported for measures of resting cardiac function, which somewhat questions the sensitivity of these measures for early detection of subclinical myocardial injury.

### Strength and limitations

A major strength of our study was the detailed assessment of PA, which included the analysis of both, a PA diary and objectively measured PA with an activity tracker, including supervised and unsupervised ET and PA. Further strengths include the randomized design and use of robust and established primary and secondary endpoints.

A major limitation is selection bias, with studies on exercise interventions often resulting in the recruitment of physically active patients. Our cohort fulfilled the WHO PA guidelines^26^ even during the AC-cycles, and the fact that 5 patients performed the ET during AC despite having been randomised to EXpostAC versus only one patient changing groups in the other direction suggests that patients were interested in completing ET as soon as possible. Furthermore, simple use of PA trackers has been shown to increase PA and steps,^48,49^ however, in cancer patients additional benefits of PA counselling or incentives has been found in few small studies.^50,51^ Against this background, our supervised ET intervention did not lead to a difference of PA between groups.

In addition, due to recruitment difficulties related to the Covid-19 pandemic, the study was limited by the small sample size (calculated sample size was 102 patients) and patients with available primary outcome data. However, based on the present results of our ITT analysis, instead of the assumed effect size of 0.6, we only achieved an effect size of 0.2, therefore we would have still been largely underpowered even if we reached target sample size. Additionally, in PP-analysis, there was a trend for the group x time interaction for GLS at AC-end (p=0.09), suggesting that GLS deteriorated more in the EXduringAC group, opposing our hypothesis. Nevertheless, due to the small sample size, results from this analysis have to be interpreted with caution. Moreover, Covid-19 related adaptations of the centre-based ET-programmes with inclusion of home-based training sessions and differences in supervised ET at the three included centres resulted in poorly standardized centre-based ET. Finally, Fitbit Zip did not provide information on the thresholds used for calculating minutes of medium and high intensity, which is another limitation of the study. The only information we received was that the 10-min criteria for minimal bout duration was used ^52^, which leads to much lower daily MVPA durations than without the 10-min bout criteria as has been recommended by the WHO since 2020.^,26,53^ Therefore, comparisons between studies are only valid for the same MVPA definitions. Our study cohort was rather young (median age 47 years [38, 57 years]) and relatively healthy, consequently a low incidence of AC-associated cardiotoxicity was found. Last but not least, study results may not be generalizable to other cancers and cardiotoxic therapies.

## Conclusion

In this RCT in early breast cancer and lymphoma patients, we found that structured ET was not superior to PA counselling and tracking on mitigation of cardiotoxicity. The assessment of objectively measured PA allowed us to demonstrate, for the first time in humans, that in our pooled and physically active cohort the dose of PA was inversely related to changes in GLS and hsTnT, suggesting that higher levels of PA may attenuate myocardial injury.

Future studies in larger patient populations are needed to confirm the results of our study, including long-term assessments to evaluate whether cardioprotective effects during AC translate into attenuation of LV dysfunction and reduced heart failure incidence.

## Data Availability

The data underlying this article will be shared upon reasonable request to the corresponding author.

## List of non-standard abbreviations

AC: anthracycline-based chemotherapies
BP: Blood pressure
CV: Cardiovascular
CVD: Cardiovascular Disease
CRF: Cardiorespiratory fitness
CORE: Cardio-oncologic rehabilitation programme
CPET: Cardiopulmonary exercise test
EF: Ejection fraction
ET: exercise training
GLS: Global longitudinal strain
hs-CRP: high-sensitivity C-Reactive Protein
hsTNT: high-sensitive Troponin-T
LAVI: left atrial end-systolic volume index
LV: Left ventricular
LVEDV: Left ventricular end-diastolic volume
LVEDVi: Left ventricular end-diastolic volume index
LVESV: Left ventricular end-systolic volume
MVPA: Moderate to vigorous physical activity
NT-proBNP: N-terminal pro-brain natriuretic peptide
PA: Physical activity
QOL: Quality of life

## Acknowledgements

We gladly acknowledge all oncologists and breast care nurses for their contribution in the recruitment of breast cancer and lymphoma patients, specifically Dr. Barbara Zeyen, Dr. Urban Novak, Monika Biedermann and Karin Biedermann from the Inselspital in Bern, Dr. Christa Baumann and Dr. Alexander Heini from the Lindenhofspital Bern, Dr. Andreas Barth and Dr. Michele Ciriolo from the Bürgerspital Solothurn and Dr. Jean-Marc Lüthi, Dr. Christoph Ackermann and Dr. Jeanne Godau from the Spital Thun. In addition, we would like to thank Dr. Konstantinos Koskinas for the evaluation of the serious adverse event in his role as the independent data safety committee of the study.

## Authors’ contributions

PE and MW were involved in the conception and design. CS and PE were involved in data acquisition and analysis of the study. CS drafted the manuscript. MW and PE were involved in the methodology and interpretation of data and revised the manuscript. KC and TS revised the manuscript critically to provide intellectual content.

## Supplemental Material

Supplemental methods

Tables S1-S4

Figure S1: Study flow illustrating patients included in per-protocol analysis.

Figure S2: Lineplots illustrating individual changes in GLS, LVEDVi, hsTnT and NT-proBNP from baseline to AC-end and follow-up.

AC, anthracycline-based chemotherapies; GLS, global longitudinal strain; LVEDVi, left-ventricular end-diastolic volume index; hsTnT, high-sensitivity Troponin-T; NT-proBNP, N-terminal pro-brain natriuretic peptide

